# Feasibility and pilot efficacy of self-applied home-based cognitive training and brain stimulation

**DOI:** 10.1101/2024.09.06.24313172

**Authors:** Merle Rocke, Anna E. Fromm, Nora Jansen, Friederike Thams, Catalina Trujillo-Llano, Ulrike Grittner, Daria Antonenko, Agnes Flöel

**Affiliations:** Department of Neurology, University Medicine Greifswald, Greifswald, Germany; Berlin Institute of Health, Charité University Medicine Berlin, Berlin, Germany; Institute of Biometry and Clinical Epidemiology, Charité University Medicine sBerlin, Corporate Member of Freie Universität Berlin, Humboldt-University Berlin and Berlin Institute of Health, Berlin, Germany; German Centre for Neurodegenerative Diseases (DZNE), Greifswald, Germany

## Abstract

**Objective:** To assess whether home-based, self-applied cognitive training combined with tDCS in older adults is feasible (primary), acceptable, and effective (secondary).

**Design:** Monocentric, double-blind, randomized (1:1), controlled trial conducted from May 2021 to July 2023, involving six at-home sessions and pre-, post- and follow-up assessments in the laboratory.

**Setting:** University Medicine Greifswald and participants’ homes.

**Participants:** Thirty older adults (60-80 years), randomized to anodal or sham tDCS group (n = 15 each).

**Interventions:** Six sessions at home over the course of two weeks (three per week) with training of letter updating and concurrent self-applied tDCS (1.5 mA, 20 min/30 s). Participants were thoroughly trained in self-application of the stimulation and handling of the material.

**Main outcomes measures:** The primary outcome was feasibility, operationalized by successfully performed interventional sessions per participant. Four or more out of six sessions successfully performed in at least 60% of all participants were defined for the trial to be deemed feasible. Secondary outcomes included: acceptability assessed via questionnaire and cognitive performance on training and transfer tasks.

**Results:** 29 participants successfully completed four or more out of six intervention sessions (96.7%, 95%-CI: [81.9, 100.0]), confirming the feasibility of the intervention (primary outcome). Overall satisfaction with the intervention and mean feasibility rating was high (93%, 95%-CI: [77.6 to 99.2]). Training (letter updating) task performance was superior in the target compared to the control group (β = 3.5, 95%-CI: [0.5 to 6.6], *p* = 0.037). There was no substantial difference in the transfer (N-back) task (β = -5.1, 95%-CI: [-14.6 to 4.5], *p* = 0.23).

**Conclusions:** Self-administered, home-based combination of cognitive training and tDCS is feasible and acceptable in older adults. Pre-defined secondary outcomes indicate superior cognitive enhancement of the trained function in the active stimulation group. Our study indicates that a Phase III trial is now warranted.

**Trial registration:** Prospectively registered (clinicalTrials.gov, NCT04817124).

**Summary boxes:** *What is known about this topic:*

- Effective interventions for age-related cognitive decline are not available and constitute a major unmet medical need
- Combining cognitive training with transcranial direct current stimulation (tDCS) is a safe, low-cost, and potentially effective approach for treating age-related cognitive decline, but currently used laboratory-based interventions over extended periods are time-consuming and laborious for both clinicians and patients, and thus not feasible in routine care.
- Feasibility, acceptability and efficacy of home-based, self-administered tDCS combined with cognitive training in older adults still need to be determined.

*What this study adds:*

- This study demonstrates that home-based, self-administered tDCS combined with cognitive training is feasible, acceptable and potentially effective for enhancing cognitive functions in older adults.
- Our findings pave the way for a subsequent Phase III clinical trial to provide high-level evidence for clinical efficacy of this approach.

## Introduction

Non-pharmacological interventions to enhance cognitive functions and decelerate neurodegenerative processes are urgently needed ^1^. In this context, pairing cognitive training with non-invasive brain stimulation such as transcranial direct current stimulation (tDCS) is a safe, low-cost, and potentially effective treatment option ^2^. Previously, we conducted clinical trials involving older adults without and with cognitive impairment, administering multisession cognitive training (i.e., intense practicing of working memory updating and decision-making) with anodal tDCS over left dorsolateral prefrontal cortex ^3,4^. These studies were performed in the laboratory, requiring frequent (i.e., 13) visits to the facility, thus necessitating not only high motivation, but also placing high demands on temporal availability and flexibility for patients, often challenging, especially for older adults in rural areas due to long distances and limited mobility. Moreover, limited clinical resources would not support such interventions in the population at large.

Importantly, the use of therapies in clinical routine is not only related to their effectiveness but also to their feasibility and acceptability ^5^. Thus, remotely controlled and independently self-applied tES approaches for at-home use may be a crucial step forward ^6^. It can enhance feasibility for patients, while reducing burdens on clinical resources (i.e., personnel, space), with the ultimate goal to incorporate home-based treatment approaches into routine clinical care ^7^. In particular, such interventions need to be tailored to older adults considering their abilities (i.e., in operating multiple technical devices or following complex instructions) to ensure their commitment.

We therefore conducted a monocentric, randomized, placebo-controlled clinical trial in older adults to determine the feasibility (primary), acceptability and efficacy (secondary) of a home-based independently self-applied tDCS-accompanied cognitive training ^8^. This approach has not yet been evaluated in a home environment without “real-time” supervision. We hypothesized that appropriate training and instruction will allow for at least 60 % of all participants to successfully complete at least two-thirds of all sessions, rendering the intervention feasible (pre-set criterion for primary outcome analysis).

## Methods

### Study design and participants

This is a monocentric, double-blinded, randomized controlled trial to assess the feasibility of a home-based combination of cognitive training and concurrent tDCS. All participants were older adults, right-handed, German native speakers and performed within age- and education-adjusted normative range in the CERAD-Plus Test Battery (memoryclinic.ch). No subject reported a history of neurological or psychiatric disorders or tDCS contraindications (see Table 1 for baseline characteristics). All participants underwent six training sessions over two weeks, along with pre-, post- and follow-up assessments (see Figure 1 for the study diagram). The control condition comprised the same cognitive training and concurrent sham tDCS. The study was performed at the University Medicine Greifswald and at participants’ homes. The full study protocol, detailing the methods, materials and eligibility criteria, was previously published ^8^. This study was approved by the ethics committee of the University Medicine Greifswald and pre-registered on the ClinicalTrials.gov website (Identifier: NCT04817124). According to the Declaration of Helsinki, all participants provided written consent before study inclusion. Participants were compensated with 130 € for their participation.

**Table 1.** Baseline characteristics of the study sample.

|  | All<br>*n* = 30 | Target<br>*n* = 15 | Control<br>*n* = 15 |
| --- | --- | --- | --- |
| Age (years) | 68.7 (5.1) | 68.4 (4.4) | 69.1 (5.8) |
| Sex (*n*, % female) | 19 (63.3) | 11 (73.3) | 8 (53.3) |
| Education (years) | 16.1 (2.1) | 15.6 (2.2) | 16.6 (2.0) |
| GDS | 1.2 (1.1) | 1.0 (0.9) | 1.5 (1.2) |
| Semantic fluency | 26.6 (4.9) | 26.9 (4.1) | 26.3 (5.8) |
| BNT (max. 15) | 14.8 (0.4) | 14.8 (0.4) | 14.7 (0.5) |
| MMSE (max. 30) | 29.7 (0.4) | 29.7 (0.5) | 29.8 (0.4) |
| Word list learning |  |  |  |
| Total | 23.5 (2.5) | 23.8 (2.5) | 23.2 (2.5) |
| Trial 1 (max. 10) | 6.3 (1.1) | 6.4 (1.0) | 6.1 (1.3) |
| Trial 2 (max. 10) | 8.3 (1.0) | 8.4 (1.1) | 8.1 (0.9) |

|  | All | Target | Control |
| --- | --- | --- | --- |
|  | *n* = 30 | *n* = 15 | *n* = 15 |
| Trial 3 (max. 10) | 9.0 (1.0) | 9.0 (0.9) | 8.9 (1.0) |
| Word list retrieval | 8.8 (1.2) | 9.1 (1.0) | 8.5 (1.4) |
| Word list intrusions | 0.0 (0.2) | 0.1 (0.3) | 0.0 (0.0) |
| Figure copying (max. 11) | 10.9 (0.3) | 11.0 (0.0) | 10.9 (0.4) |
| Figure retrieval (max. 11) | 10.3 (1.2) | 10.3 (1.2) | 10.2 (1.1) |
| Phonematic fluency | 15.5 (3.7) | 15.8 (4.3) | 15.1 (3.1) |
| Trial-making test |  |  |  |
| Part A (sec) | 36.7 (10.7) | 36.5 (9.7) | 36.8 (12.0) |
| Part B (sec) | 70.6 (17.1) | 68.0 (14.5) | 73.1 (19.6) |
| Digit-span |  |  |  |
| Forward | 7.8 (1.9) | 7.8 (1.7) | 7.9 (2.1) |
| Backward | 6.2 (1.8) | 6.7 (1.7) | 5.8 (1.9) |

Data are shown as the mean (SD) or *n*. GDS, Geriatric Depression Scale. BNT, Boston Naming Test. MMSE, Mini Mental Status Examination.

**Figure 1.**
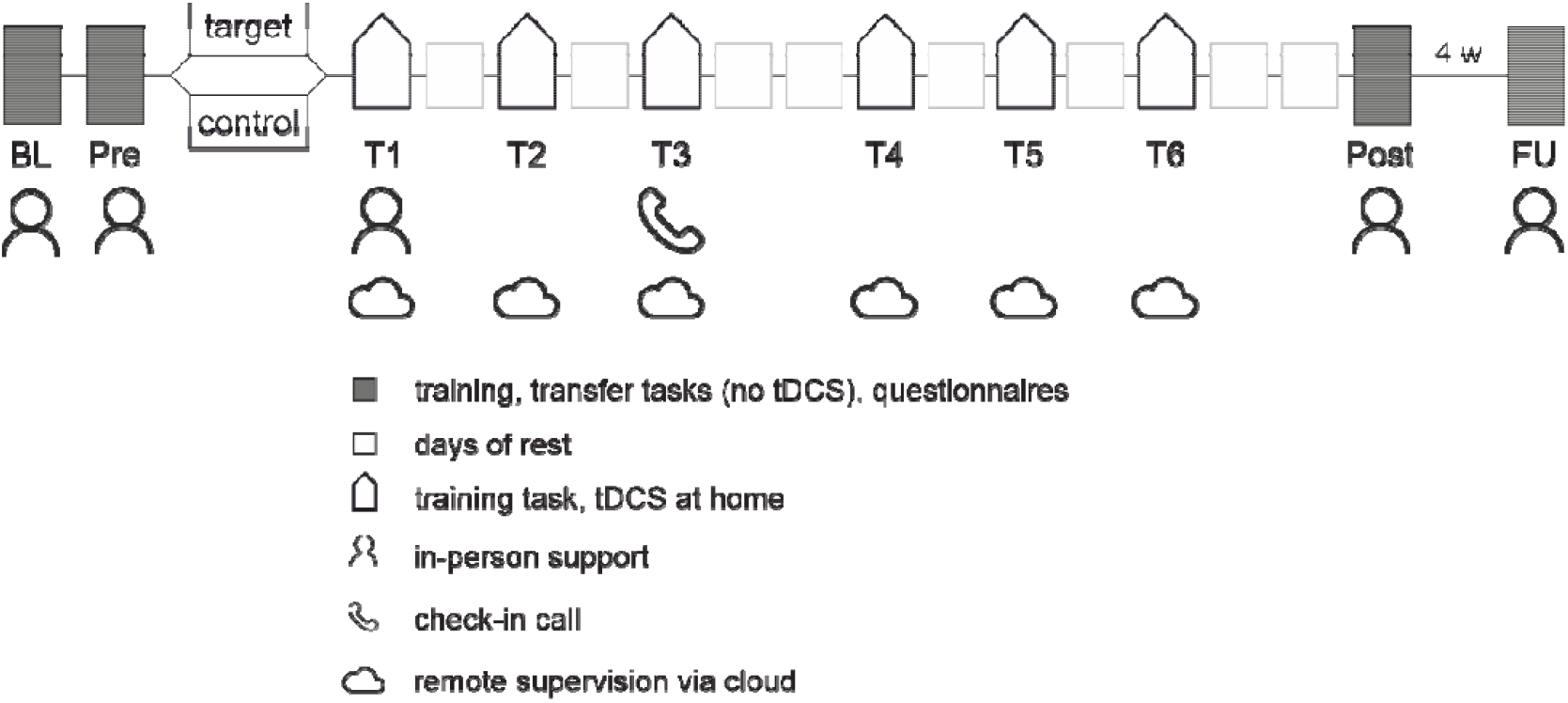
Study diagram. During the training period the study team was always available via telephone. At pre-assessment and T1, the focus was on training participants for self-application. Training sessions were conducted three times a week, with a rest day between sessions and no weekend sessions. tDCS, transcranial direct current stimulation. BL, baseline assessment. Pre, pre-assessment. T, training session. FU, follow-up assessment.

### Randomization and blinding

Thirty participants were randomly allocated in a 1:1 ratio to the target intervention group or the control group based on their baseline performance in the letter updating task (LU). To ensure blinding, the control group received sham stimulation, where a current was applied for 20 seconds at the beginning of the stimulation to mimic the tDCS tingling sensation and then gradually reduced to zero over the next 20 seconds. Previous studies have validated sham stimulation as an effective blinding method, showing no additional adverse effects compared to anodal stimulation ^9^. Additionally, at the end of all training sessions, participants were asked to guess their assigned treatment group and blinding success was assessed through the James Blinding Index (BI, see below).

To ensure investigators’ blinding, the stimulation protocols were encoded, and another researcher, who was not involved in data acquisition, handled participant allocation ^8^.

### Intervention

After randomization at baseline assessment, participants attended a pre-assessment, followed by two weeks of training sessions (three training sessions per week, every other day excluding weekends, totaling six sessions), and a post-assessment. A follow-up assessment was conducted one month after the post-assessment (see Figure 1). All training sessions were conducted at participants’ homes. The first session was supervised in person by study staff, while subsequent sessions were self-applied and remotely supervised.

In each of the six training sessions, participants performed a well-established LU task ^10,11^ on a tablet computer. Lists of letters A to D, with varying lengths (5, 7, 9, 11, 13 or 15 letters; six times each, totaling 36 lists), were presented in random order, one letter at a time (presentation duration: 2000 ms; ISI: 500 ms). After each list, participants were asked to recall the last four presented letters in their respective order. Simultaneously, participants self-applied either anodal or sham tDCS using a battery-operated stimulator (Starstim-Home Research Kit, Neuroelectrics, Spain). Participants mounted two round saline-soaked sponge electrodes in a neoprene head cap at the beginning of each session. The anodal and cathodal electrodes were placed over the left dorsolateral prefrontal cortex (F3) and the right supraorbital cortex (Fp2), according to the 10-20 EEG grid system, respectively. For anodal tDCS, a 20-second ramp-up period was followed by 20 minutes at 1.5 mA and then a 20-second ramp-down. Sham tDCS involved only the ramp-up and ramp-down, totaling 40 seconds of stimulation, with the cap remaining on for the entire session. Adverse events were assessed via a questionnaire during the third and sixth sessions. Participant progress was monitored remotely via the cloud, with telephone support and remote access to the tablet computer available as needed.

During the pre-, post- and follow-up assessments, participants performed both the training (LU) and transfer (numerical N-back) tasks. The numerical N-back task, used to assess transfer to an untrained working memory task, included 1-back and 2-back conditions and was performed on a laptop. Each condition consisted of 9 trials and 10 items (presentation duration 1500 ms, ISI 2500 ms). Participants were asked to compare each presented number to the one ‘n’ steps back and determine if they were identical. The pre-assessment also included comprehensive training on the stimulation setup. At the post-assessment, participants completed a feasibility questionnaire. Baseline assessment included a complete neuropsychological evaluation and supervised training of the intervention along with the training task (LU). For a more comprehensive overview, please refer to Thams et al. ^8^.

### Outcomes

The primary outcome measure was the feasibility of home-based tDCS and cognitive training, defined as each participant successfully completing at least two-thirds of the intervention sessions (≥ 4 out of 6 sessions). A session was considered successful if it was recorded as completed in the cloud and the participant did not contact the study staff to report issues or request rescheduling. The a priori set feasibility criterion was that at least 60% of all participants met this threshold. Secondary outcomes included acceptability, measured by a self-reported questionnaire ^8,12^, and working memory performance in trained and untrained tasks assessed at post- and follow-up, operationalized by the number of correctly responded lists in the LU task and the percentage of correct answers in the N-back task, respectively.

### Statistical analysis

Predefined statistical analyses were conducted using R ^13^ (version 4.3.1), described in the statistical analysis plan uploaded before any analysis was conducted. All participants were included in the full dataset. Analysis of the primary outcome was based on descriptive statistics. The point estimate of the feasibility criterion and the corresponding 95 %-CI was calculated. Separate linear mixed models were conducted for post-assessment and follow-up timepoints, for each task (LU and N-back). Model-based marginal means, group differences and 95 %-CI are reported ^14^. A two-sided significance level of 0.05 was used.

Safety outcomes are reported as incidences (*n*, incidence rate with 95%-CI) overall and by intervention group, based on the safety analysis set. An adverse event was defined as at least a moderate stimulation sensation. Incidence rates and 95%-CI were calculated using Poisson regression models, accounting for different observation periods for each participant. Group comparisons were conducted using incidence rate ratios and 95%-CI. Blinding success was measured through the BI, which ranges from 0 (indicating no blinding with all answers correct) to 1 (indicating no blinding with all answers incorrect). A value of 0.5 indicates random guessing.

## Results

From March 2021 to July 2023, we screened 123 potential participants via telephone. Of these, 55 participants were invited to on-site screening and baseline assessment. After excluding 93 participants, 30 were included in the trial and randomly assigned to either active (target intervention, *n* = 15) or sham tDCS (control intervention group, *n* = 15) combined with cognitive training (see Supplementary Figure 1 for the CONSORT diagram). The last post-assessment was completed on Oct. 2^nd^, 2023, with the last follow-up assessment following on Nov 1^st^, 2023. No participants dropped out, resulting in 30 participants for the analyses (mean/SD age 69 ± 5 years, 19 women).

### Feasibility

Out of 30 enrolled participants, 29 participants completed four or more of the six intervention sessions successfully, which amounts to 96.7% (95%-CI: [81.9 to 100.0]), confirming that the intervention was feasible (primary outcome, see Supplementary Figure 2 for a more detailed overview of completed sessions). On the acceptability questionnaire (secondary outcome), the item that assessed overall satisfaction with the intervention was rated “agree” (*n* = 10) or “strongly agree” (*n* = 18) by 93% of participants (*n* = 28). Overall, mean for acceptability (i.e., satisfaction and feasibility) ratings was 93% (95%-CI: [77.6 to 99.2]) on a 5-point Likert scale (see Figure 2). Most of the items of the questionnaire were rated high by the majority of participants (i.e., confidence in setting up the system: 93%, comfort with tDCS: 83%).

**Figure 2.**
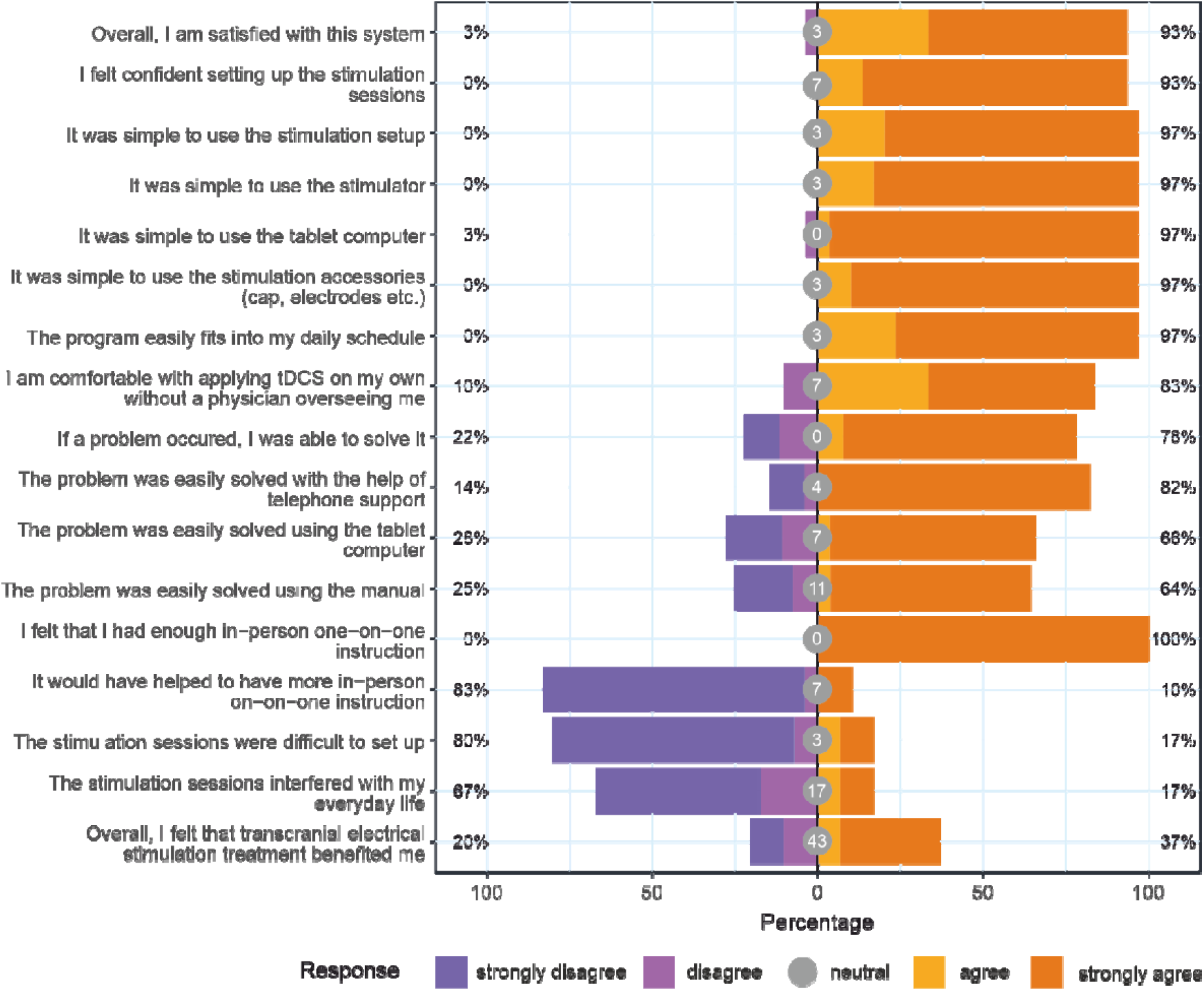
Acceptability questionnaire. Responses to all items by all participants in percent.

### Cognitive effects

#### Training task

The overall training task (LU) performance was superior in the target compared to the control group (β = 3.5, 95%-CI: [0.5 to 6.6], *p* = 0.037; model-derived marginal means: 18.9, 95%-CI: [16.7 to 21.1] for target and 15.7, 95%-CI: [13.6 to 17.8] for control group; see Figure 3A). No substantial immediate training effect emerged at the post-appointment (β = 2.8, 95%-CI: [−0.5 to 6.2], *p* = 0.096, model-derived marginal means: 20.6, 95%-CI: [18.2 to 23.0] for anodal and 17.8, 95%-CI: [15.5 to 20.0] for sham group).

**Figure 3.**
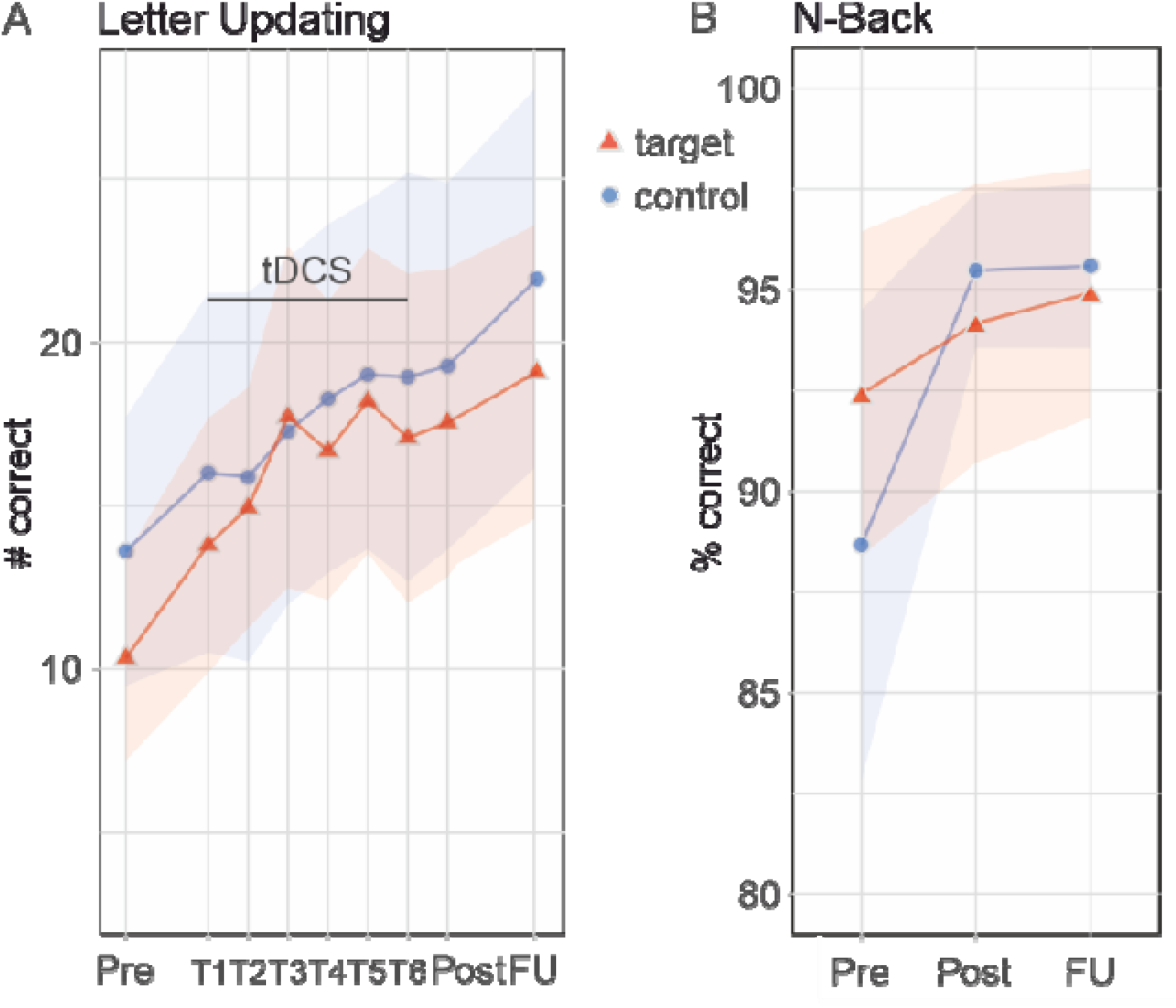
Performance on training and transfer tasks. (A) Overall improvement over the intervention period in letter updating training task. Task performance was superior in the target compared to the control group (β = 3.5, 95%-CI: [0.52 to 6.58], *p* = 0.037 (model-derived marginal means: 18.9, 95%-CI: [16.7 to 21.1] for target and 15.7, 95%-CI: [13.6 to 17.8] for sham control group; *n* = 30 participants, 206 data points). (B) N-back transfer task. Performance did not differ between groups (β = -5.1, 95%-CI: [-14.6 to 4.5], *p* = 0.23 (model-derived marginal means: 80.5, 95%-CI: [73.7 to 87.2] for the target group and 85.5, 95%-CI: [79.2 to 91.8] for the sham control group; *n* = 30 participants, 60 data points). tDCS, transcranial direct current stimulation. Pre, pre-assessment. FU, follow-up assessment (one month after the intervention).

Explorative analyses revealed a substantial treatment effect after the first week of training (β = 3.3, 95%-CI: [0.2, 6.5], *p* = 0.04, model-derived 20.7, 95%-CI: [17.6 to 23.9] for target and 19.6, 95%-CI: [16.6 to 22.6] for control group). At the follow-up time point, we observed no substantial treatment effect (β = 1.4, 95%-CI: [−2.3 to 5.2], *p* = 0.45, model-derived marginal means: 21.6, 95%-CI: [18.9 to 24.3] for target and 20.1, 95%-CI: [17.6 to 22.7] for control group).

#### Transfer task

Transfer task (N-back) performance did not differ substantially between groups at the post-appointment (β = -5.1, 95%-CI: [−14.6 to 4.5], *p* = 0.23; model-derived marginal means: 80.5, 95%-CI: [73.7 to 87.2] for target and 85.5, 95%-CI: [79.2 to 91.8] for control group; see Figure 3B). At the follow-up time point, we observed no substantial treatment effect as well (β = -1.9, 95%-CI: [−9.9 to 6.0], *p* = 0.61, model-derived marginal means: 83.1, 95%-CI: [77.5 to 88.7] for target and 85.1, 95%-CI: [79.9 to 90.3] for control group).

### Adverse events

Two participants in the target group reported four adverse events (itching and heating), while no adverse events were reported in the control group. No serious adverse events were reported, and no participant terminated participation due to adverse events (see Supplementary Table 1).

### Blinding

The BI estimate was 0.7 (95%-CI: 0.5 to 0.8), indicating that participants were effectively blinded (see Supplementary Table 2).

## Discussion

Our findings demonstrate excellent feasibility and acceptability of the combined self-administered at-home intervention in older adults. The prespecified criterion for inference of feasibility was exceeded. Only one participant did not complete all sessions, thus overall commitment and retention rates were very high, with no dropouts or losses to follow-up. A questionnaire further showed overall high acceptability, with excellent scores for tolerability and satisfaction with the intervention and very low discomfort ratings, with only two participants reporting minor side effects. To the best of our knowledge, this is the first Phase II clinical trial of self-administered approach of cognitive training combined with tDCS in a cohort of older adults ^14^. Combined approaches challenge participants in terms of assembling study material, such as inserting electrodes into the cap, putting on the cap, switching on the stimulator, checking electrode impedances, as well as starting the training session on the tablet computer, a concern particularly valid for older adults who are often less experienced in handling technical devices and software ^15^. Our findings confirm an exploratory analysis of five older participants (however of “younger old” age, i.e., 51-68 years) who performed a motor training together with tDCS on their own at home ^16^. Our participants particularly appreciated detailed guidance and training on the practical aspect of this approach. Given that no “real-time” or video supervision is needed for this approach, demands on personnel and laboratory space are reduced for the investigator. Moreover, the at-home approach reduces time and travel commitment for participants ^17^, and may thus overcome barriers to neuromodulation as a treatment for routine clinical use ^15,18^.

Secondary analyses on efficacy of active stimulation revealed superior training (LU), but no robust effects for transfer task performance (N-back). Overall, the behavioral results were in line with beneficial effects of the combined approach for cognitive outcomes in our previous clinical trials conducted in the laboratory in healthy older adults ^4^ and older adults with cognitive impairment ^3^. However, while these trials showed group differences for transfer working memory (N-back) task performance only, we observed an effect of tDCS on the trained (LU) task performance. The training task effect in the present study may be related to higher stimulation intensity (1.5 instead of 1 mA). With regard to the lack of transfer effects, we can only speculate that more concurrent training and stimulation sessions may be needed (i.e., nine ^3,4^ instead of six sessions in the present report). Moreover, our previous studies included an additional decision-making training in each session, which may have contributed to the transfer effects.

### Strengths and limitations of this study

The TrainStim-Home Trial was co-designed with the input of five healthy older adults who had previously participated in a similar laboratory-based study ^4^. These participants were invited to assess the trial procedures and training materials, including the manuals for handling stimulation devices and tablet computers. Their extensive feedback was carefully integrated into the program. For instance, the trial sessions highlighted challenges, such as multi-stage instructions for set-up of stimulation equipment being too difficult and confusing to follow for participants. To address this, we simplified the set-up, and developed additional aids, including a checklist and an instruction manual with additional printed as well as video-based visual aids. The 20-minute video was then shown to all participants during the baseline assessment and remained accessible throughout the treatment period as an on-demand resource on their tablet computers ^6^. With these improvements, participants were able to confidently and accurately set up the stimulation equipment. Our comprehensive manual provided standardized and reproducible set-up and delivering of stimulation by participants, thus ensuring quality and safety. By delivering the intervention at home, we were able to increase accessibility for participants from rural areas who are disadvantaged due to long distances to the University in rural areas, or participants with concomitant deficits in ambulation.

Limitations include our rather small cohort as well as training regimen over two weeks only, issues to be addressed now in a large multicenter Phase III trial.

## Conclusion

In conclusion, feasibility and acceptability of self-administered cognitive training combined with tDCS in older adults were clearly demonstrated, with indications for efficacy in terms of cognitive outcome in an at-home approach, which shows significant advantages for both investigators and patients. These findings warrant a subsequent Phase III trial, which would, if successful, introduce a safe and feasible approach to prevent cognitive decline into clinical practice.

## Data Availability

All data produced in the present study are available upon reasonable request to the authors.

## Author contributions

DA and AF designed the study. MR, FT, and NJ collected the data. MR, AEF, FT, CTLL, UG, and DA performed data analyses. MR, DA and AF wrote the manuscript. All authors reviewed and approved the final version of the manuscript.

## Ethical approval

In accordance with the Declaration of Helsinki, all participants provided signed informed consent before participating in the study. The ethics committee of University Medicine Greifswald approved this study (ID: BB002121).

## Transparency statement

The lead authors affirm that the manuscript is an honest, accurate, and transparent account of the study being reported, that no important aspects of the study have been omitted and that any discrepancies from the study as originally planned have been explained.

## Reporting on sex and gender

This research applies to both sexes, as it includes both female (*n* = 19) and male (*n* = 11) participants. All analyses were controlled for the effects of age and self-reported sex. Gender-related factors were not considered in the data analyses.

## Data sharing

The data supporting this study’s findings are available from the corresponding authors upon reasonable request.

