## Supplementary Material for "Feasibility and pilot efficacy of self-applied home-based cognitive training and brain stimulation"

### Supplementary figures

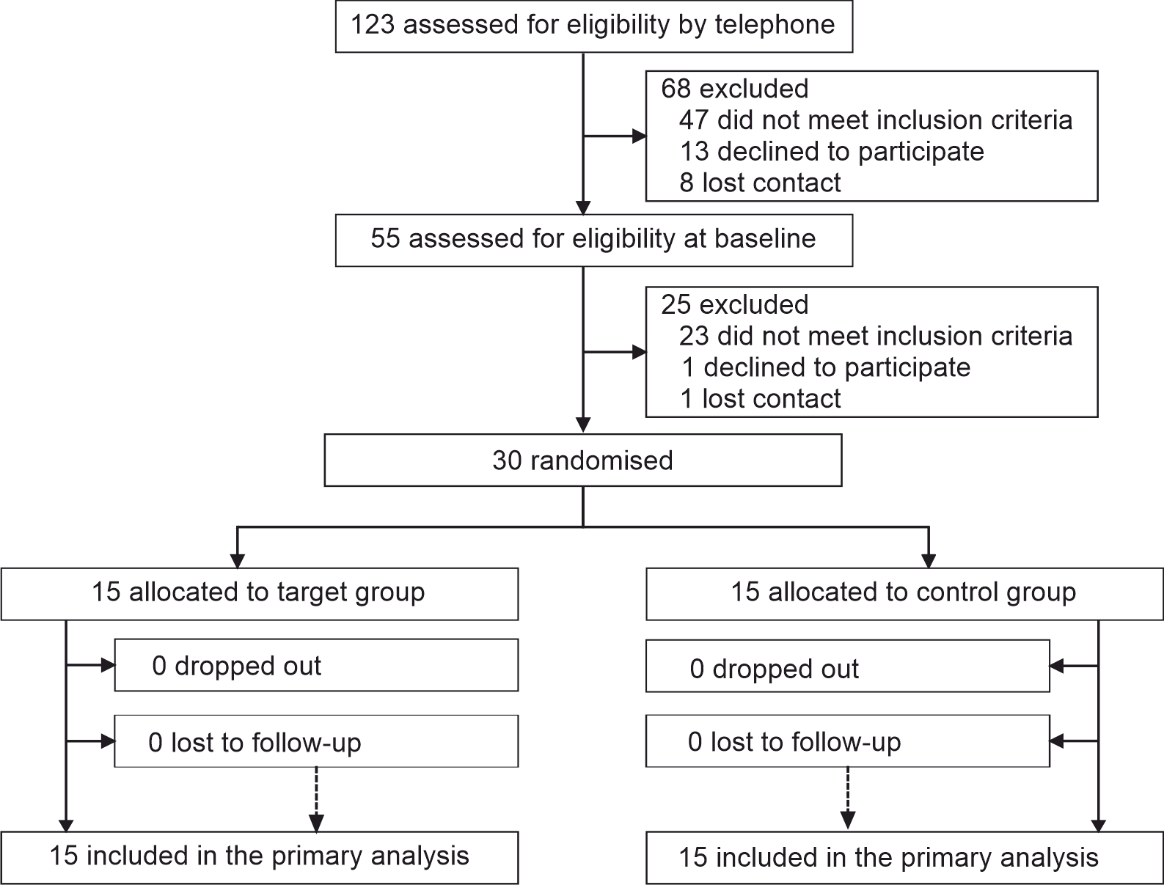

**Supplementary Figure 1.** CONSORT diagram.

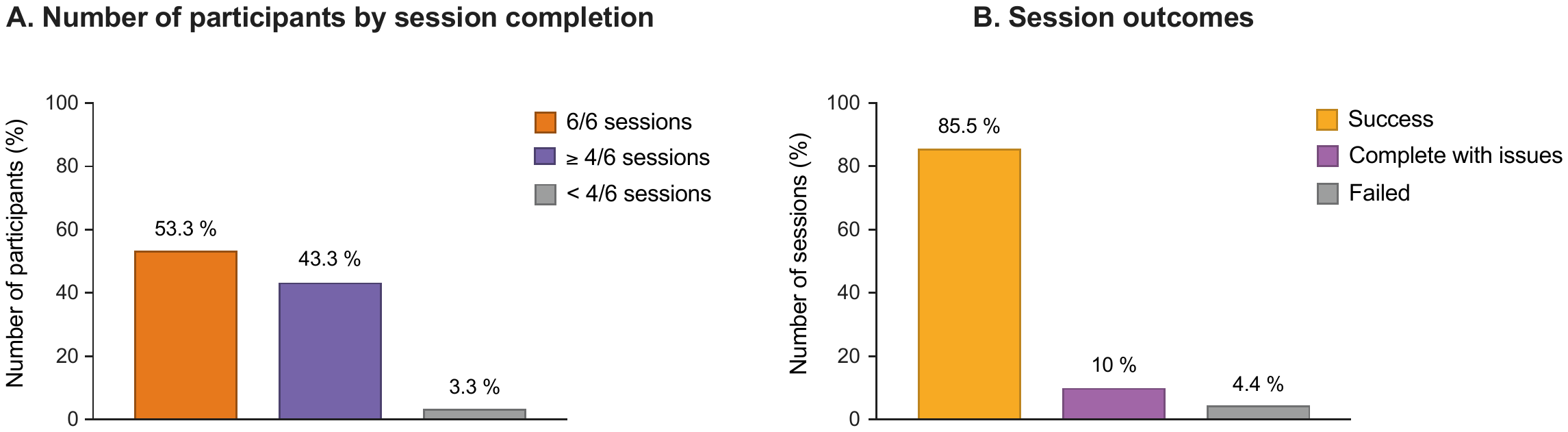

**Supplementary Figure 2.** Participant session completion overview. (A) Number of participants by session completion. The bar plot shows the percentage of participants based on the number of sessions they completed. A session was considered successful if it was recorded as completed in the cloud and the participant had not contacted the study staff to report issues or request rescheduling. Among the participants, 16 (53.3%) completed all 6 sessions, 13 (43.3%) completed 4 or 5 sessions, and 1 (3.3%) completed fewer than 4 sessions. Thus, 29 participants met the feasibility criterion. One participant experienced technical issues (abort of the stimulation due to high impedance and bad Wi-Fi connection and, thus, incomplete synchronization with the cloud server). (B) Session outcomes. The bar plot displays the percentage of sessions based on their outcomes. Of all sessions, 154 (85.5%) were successful, 18 (10%) were completed but had reported issues that were resolved, and 8 (4.4%) failed due to unsolvable problems (i.e., system abortion, scheduling issues).

### Supplementary tables

| **Supplementary Table 1**. Self-reported incidence of adverse events (at least moderate symptoms) by group during intervention. | | | | | |
| --- | --- | --- | --- | --- | --- |
|  | | **All**  *n* = 30 | **Target**  *n* = 15 | **Control**  *n* = 15 | **Incidence rate ratio for group differences**  (95 %-CI) |
| Observation time in days, mean (SD) | | 6 (0) | 6 (0) | 6 (0) |  |
| Total number of adverse events | | 4 / 0 (0 to 0.1) | 4 / 0 (0 to 0.1) | 0 / .. | .. |
|  | Itching | 2 / 0.01 (0 to 0.03) | 2 / 0.02 (0 to 0.07) | 0 / .. | .. |
|  | Pain | 0 / .. | 0 / .. | 0 / .. | .. |
|  | Burning | 0 / .. | 0 / .. | 0 / .. | .. |
|  | Warmth/heat | 1 / 0.01 (0 to 0.02) | 1 / 0.01 (0 to 0.05) | 0 / .. | .. |
|  | Metallic/iron taste | 0 / .. | 0 / .. | 0 / .. | .. |
|  | Fatigue/decreased alertness | 1 / 0.01 (0 to 0.02) | 1 / 0.01 (0 to 0.05) | 0 / .. | .. |
|  | Other | 0 / .. | 0 / .. | 0 / .. | .. |
| Reported values are absolute frequency of the respective adverse events / incidence rate (95 %-CI). | | | | | |

| **Supplementary Table 2.** Number of participants by group assignment and guess | | | | | |
| --- | --- | --- | --- | --- | --- |
| **Assignment** |  | **Response** | | | |
|  |  | Treatment | Control | DK | Total |
|  | Treatment | 4 | 7 | 4 | 15 |
|  | Control | 3 | 8 | 4 | 15 |
|  | Total | 7 | 15 | 8 | 30 |
| DK, “Don’t know”. | | | | | |
